# COVID-19 vaccine antibody response is associated with side-effects, chronic health conditions, and vaccine type in a large Northern California cohort

**DOI:** 10.1101/2022.09.30.22280166

**Authors:** Olivia Solomon, Cameron Adams, Mary Horton, Marcus P. Wong, Michelle Meas, Xiaorong Shao, Indro Fedrigo, Samantha Hernandez, Hong L. Quach, Diana L. Quach, Anna L. Barcellos, Josefina Coloma, Michael Busch, Eva Harris, Lisa F. Barcellos

**Author notes:** **Corresponding author:** Lisa F. Barcellos, 324 Stanley Hall, Berkeley, CA 94720.

## Abstract

As vaccines have become available for COVID-19, it is important to understand factors that may impact response. The objective of this study is to describe vaccine response in a well-characterized Northern California cohort, including differences in side-effects and antibody response by vaccine type, sex, and age, as well as describe responses in subjects with pre-existing health conditions that are known risk factors for more severe COVID-19 infection. From July 2020 to March 2021, ∼5,500 adults from the East Bay Area in Northern California were followed as part of a longitudinal cohort study. Comprehensive questionnaire data and biospecimens for COVID-19 antibody testing were collected at multiple time-points. All subjects were at least 18 years of age and members of the East-Bay COVID-19 cohort who answered questionnaires related to vaccination status and side-effects at two time-points. Three vaccines, Moderna (2 doses), Pfizer-BioNTech (2 doses), and Johnson & Johnson (single dose), were examined as exposures. Additionally, pre-existing health conditions were assessed. The main outcomes of interest were anti-SARS-CoV-2 Spike antibody response (measured by S/C ratio in the Ortho VITROS assay) and self-reporting of 11 potential vaccine side effects. When comparing both doses of the Moderna vaccine to respective doses of Pfizer-BioNTech, participants receiving the Moderna vaccine had higher odds of many reported side-effects. The same was true comparing the single-dose Johnson & Johnson vaccine to dose 2 of the Pfizer-BioNTech vaccine. The antibody S/C ratio also increased with each additional side-effect after the second dose. S/C ratios after vaccination were lower in participants aged 65 and older, and higher in females. At all vaccination timepoints, Moderna vaccine recipients had a higher S/C ratio. Individuals who were fully vaccinated with Pfizer-BioNTech had a 72.4% lower S/C ratio compared to those who were fully vaccinated with Moderna. Subjects with asthma, diabetes, and cardiovascular disease all demonstrated more than a 20% decrease in S/C ratio. In support of previous findings, we show that antibody response to the Moderna vaccine is higher than the Pfizer-BioNTech vaccine. We also observed that antibody response was associated with side-effects, and participants with a history of asthma, diabetes, and cardiovascular disease had lower antibody responses. This information is important to consider as further vaccines are recommended.

## Introduction

By February 2021, three COVID-19 vaccines had received emergency use authorization; the two-dose Pfizer-BioNTech, followed by the two-dose Moderna, and finally the single-dose Johnson and Johnson vaccination. All three vaccines have been shown to be effective at reducing SARS-CoV-2 infection and severity/hospitalization of COVID-19^1-3^. In the United States, all three vaccines were widely administered following authorization, and in Europe, the two-dose AstraZeneca vaccine was also dispersed.

With multiple successful vaccines being administered world-wide, the identification of side-effects and antibody response in different age groups and between males and females was of interest. The largest studies were released from the United Kingdom, including comparisons primarily between the AstraZeneca and Pfizer vaccines. In a study of 45,965 UK residents, Wei et al.^4^ reported that both AstraZeneca and Pfizer vaccines generated a lower antibody response after the first dose in older subjects >60 years of age compared to those <60 years of age but that after the second dose, the response had equalized between the age groups. The study additionally reported a lower antibody response in males compared to females. An investigation of SYNLAB Estonia employees (n = 122) by Naaber et al.^5^ also reported a lower antibody response in older patients along with a lower number of side-effects reported by older patients for the Pfizer vaccine.

Of particular interest for the United States was the comparison of Pfizer and Moderna vaccines, as these were the most widely available options. Studies have consistently shown that the Moderna vaccine produces a higher antibody response compared to the Pfizer vaccine. A small study in adults affiliated with the University of Virginia (n = 167) reported lower antibody response in those over age 50 comparted to those under 50, with an overall higher response to the Moderna vaccine compared to Pfizer^6^. Brockman et al.^7^ reported a lower and more quickly declining humoral response in older vaccine recipients compared to younger. In 215 healthy participants in Boston, Naranbhai et al^8^. showed a higher antibody response in recipients of the Moderna vaccine compared to the Pfizer vaccine and reported that both of the two-dose mRNA vaccines had a higher antibody response than the single dose J&J vaccine. In another sample of 289 subjects from Quatar University, which included both naïve and previously infected subjects, authors reported higher antibody levels in those who received the Moderna compared to the Pfizer vaccine^9^. In 1,674 healthcare workers in Belgium, another study by Steensels et al.^10^ similarly concluded there is a higher antibody response from Moderna compared to Pfizer.

In addition to antibody response in relatively healthy individuals, the immune response in immunocompromised individuals or those with pre-existing health conditions at the greatest risk of complication from COVID-19 is important to determine. A study by Embi et al.^11^ reported the effectiveness of mRNA vaccines in immunocompromised patients was 77% compared to 90% in non-immunocompromised, and another study from Deepak et al.^12^: described lower antibody titers in those who were immunocompromised. A very low antibody response was observed in patients with multiple myeloma, and it was recommended that high-risk groups with diseases such as multiple myeloma should undergo post-vaccination antibody testing^13^. Additionally, in subjects with weakened immune systems due to solid organ transplants, two doses of the mRNA vaccines did not yield a strong immune response; however, and a third dose improved this response significantly^14^.

We studied a well-characterized Northern California cohort for differences in side-effects and antibody response by vaccine type, sex, and age. We also investigated antibody responses in subjects with pre-existing health conditions that are known risk factors for severe COVID-19 infection. From July 2020 to March 2021, ∼5,500 adults from the East Bay Area in Northern California were followed. Each participant provided comprehensive questionnaire data and provided biospecimens for COVID-19 antibody testing. In the current study, we focused on two timepoints, defined as timepoint 1 and timepoint 2. Timepoint 1 captured early vaccinations for this cohort and biospecimens were collected. For timepoint 2, additional data on vaccination side effects were collected.

## Methods

### Study design and participants

All subjects were participants in the East Bay COVID-19 cohort who completed timepoint 1 (February – March 2021) or timepoint 2 (May – June 2021) study questionnaires. The original study design has been previously described^15^. At both timepoint 1 and timepoint 2, the study questionnaire collected detailed information on vaccination, including vaccination status, date(s) of vaccination, side-effects resulting from vaccination, and pre-existing health conditions. Additionally, at timepoint 1, Ortho VITROS antibody S/C ratio was measured; blood sample collection and laboratory methods have been previously described^16^. A summary of study subject characteristics is provided in Table 1.

**Table 1.** Demographics and other characteristics of study participants.

|  | **Timepoint 1**<br>**All Subjects**<br>**(n=4,714)** | **Timepoint 1**<br>**Analysis**<br>**subset<sup>a</sup>**<br>**(n=2,048)** | **Timepoint 2**<br>**All Subjects**<br>**(n=4,642)** | **Timepoint 2**<br>**Analysis**<br>**subset<sup>b</sup>**<br>**(n=4,423)** |
| --- | --- | --- | --- | --- |
| **Age, mean (SD)** | 52.67 (15.21) | 60.36 (14.92) | 52.60 (15.21) | 52.57 (15.14) |
| **Age No. (%)** |  |  |  |  |
| 18-29 | 279 (5.9) | 65 (3.2) | 276 (6.0) | 265 (6.0) |
| 30-44 | 1292 (27.5) | 338 (16.5) | 1290 (27.8) | 1221 (27.6) |
| 45-64 | 1854 (39.5) | 535 (26.1) | 1827 (39.4) | 1755 (39.7) |
| 65-74 | 966 (20.6) | 830 (40.5) | 941 (20.3) | 900 (20.4) |
| ≥75 | 307 (6.5) | 280 (13.7) | 303 (6.5) | 277 (6.3) |
| **Race/Ethnicity No. (%)** |  |  |  |  |
| African American | 118 (2.8) | 56 (3.1) | 140 (3.0) | 130 (2.9) |
| AMI | 67 (1.6) | 21 (1.1) | 85 (1.8) | 80 (1.8) |
| Asian/Pacific Islander | 586 (13.9) | 198 (10.8) | 635 (13.7) | 608 (13.8) |
| Hispanic | 421 (10.0) | 145 (7.9) | 488 (10.5) | 457 (10.3) |
| Two or more | 282 (6.7) | 69 (3.8) | 320 (6.9) | 310 (7.0) |
| White | 2728 (64.9) | 1342 (73.3) | 2968 (64.0) | 2832 (64.1) |
| **Education No. (%)** |  |  |  |  |
| < College | 334 (7.9) | 145 (7.9) | 381 (8.2) | 349 (7.9) |
| ≥Completed College | 3872 (92.1) | 1686 (92.1) | 4257 (91.8) | 4070 (92.1) |
| **Sex No. (%)** |  |  |  |  |
| Male | 1497 (35.6) | 589 (32.2) | 1658 (35.7) | 1580 (35.7) |
| Female | 2711 (64.4) | 1243 (67.8) | 2983 (64.3) | 2842 (64.3) |
| **Vaccine Type No. (%)** |  |  |  |  |
| Johnson & Johnson | 1 (0.0) | - | 291 (6.3) | 281 (6.4) |
| Moderna | 950 (21.0) | 944 (46.1) | 1511 (32.6) | 1476 (33.4) |
| Pfizer-BioNtech | 1106 (24.4) | 1104 (53.9) | 2715 (58.7) | 2666 (60.3) |
| Astrazeneca | - | - | 5 (0.1) | - |
| Other | - | - | 6 (0.1) | - |
| Trial | - | - | 28 (0.6) | - |
| Unvaccinated | 2477 (54.6) | - | 72 (1.6) | - |
| **Vaccination Dose No. (%)** |  |  |  |  |
| 1<sup>st</sup> dose | 1194 (26.3) | 1184 (57.8) | 15 (0.3) | 12 (0.3) |
| 2<sup>nd</sup> dose<sup>c</sup> | 870 (19.2) | 864 (42.2) | 4555 (98.1) | 4214 (99.7) |
| Not yet vaccinated | 2477 (54.5) | - | 72 (1.6) | - |

<sup>a</sup> Vaccination with Pfizer-BioNtech or Moderna, antibody response data at timepoint 1, and record of previous infection status. Record of previous infection status means subjects have a valid antibody call on record prior to timepoint 1.
<sup>b</sup> Vaccination with Pfizer-BioNtech, Moderna, or J&J, record of previous infection status. Record of previous infection status means subjects have a valid antibody call on record prior to timepoint 1.
<sup>c</sup> Second dose includes those who received the single shot of Johnson & Johnson vaccine.

### Vaccination side effects and vaccine type

Data on a total of 11 potential vaccination side-effects for each dose of the vaccine were collected, including: pain at the injection site, rash at the injection site, fever/chills, headache, body ache, fatigue, nausea, diarrhea, abdominal pain, rash at an area other than the injection side, and redness at the injection site. In addition, study participants were asked about whether they experienced severe side effects, including hives and anaphylaxis.

Multivariate logistic regression was used to estimate the relationship between each side effect (yes/no) and vaccine type (Pfizer-BioNtech, Moderna, Johnson & Johnson). Models were adjusted for age, previous infection (defined as a reactive antibody result at timepoints prior to timepoint 1 and 2), and sex. Models were also stratified by sex to assess effect measure modification. The reported side-effects for the single-dose Johnson & Johnson vaccine were compared to the second dose of the two-dose Pfizer-BioNtech and Moderna vaccines. We additionally used Poisson regression to determine whether the count of total side-effects reported by subjects was associated with vaccine type.

### Vaccination antibody response and side-effects

To assess whether increased side-effects were associated with vaccination antibody response (measured by Ortho VITOROS S/C ratio), we first generated a subset our sample of subjects at timepoint 1 of the study who had at least 14 days between their last vaccination dose and the collection of their blood sample. We used multivariable linear regression to estimate the relationship between side-effects (yes/no) and antibody S/C ratio, adjusting for age, sex, number of days between vaccination and blood draw, and vaccine type. Analyses were stratified by those who were 14 days past their first vaccination dose and those who were 14 days past their second vaccination dose.

### Vaccination antibody response and age and sex

In order to assess the relationship between vaccination antibody response and subject age, we first generated a subset our sample consisting of subjects at timepoint 1 of the study who reported receiving at least one dose of the COVID-19 vaccine and had a recorded antibody S/C ratio. Of the total 4,714 subjects in round three, 2,049 had received at least one vaccination dose and had an S/C ratio recorded. Since only one subject reported receiving the Johnson & Johnson vaccine by this timepoint, only the 2,048 who received Pfizer-BioNtech or Moderna were included.

We used multivariable linear regression with log transformed S/C ratio as the outcome and binary age as the predictor (<65, >=65 years old), adjusting for sex and the number of days between vaccination dose and blood draw. Analyses were stratified according to whether subjects had received only one dose of the vaccine or both doses of the vaccine by the time of blood collection.

### Vaccination antibody response and vaccine type

We also examined whether there was a relationship between antibody S/C ratio and vaccine type. We used four separate linear regression models including: (1) those who had received only dose one of the vaccination; (2) those who had received only dose one of the vaccination more than 14 days prior to their blood collection; (3) those who had received both doses of the vaccination; and (4) those who had received both doses of their vaccination 14 days or more prior to their blood collection and were considered “fully vaccinated”. All models were adjusted for age, sex, and number of days between vaccination and the time of blood collection.

### Vaccination antibody response and other health factors

Questionnaires also inquired about prior and current health conditions. For conditions with >5% frequency in our study sample, we investigated the association between S/C ratio and reported health conditions. These conditions included: asthma, pneumonia, hypertension, diabetes, cardiovascular disease, autoimmune disease, cancer, and depression. Additional factors included BMI and binge drinking (defined as >5 or >4 drinks on average when drinking in the past month for males and females, respectively). All multivariable linear regression models adjusted for vaccine type, age, sex, vaccine dose, and number of days between vaccination and blood collection.

## Results

### Vaccination demographics

Across 12 East Bay cities, vaccination rates were high and consistent across zip-codes with a median vaccination rate of 98.5% (min=93.1%, max=100%, sd=1.7%). At timepoint 2 (May – June 2021), only 72 participants (1.6%) had not received at least one dose of the vaccination. Characteristics of the unvaccinated participants can be seen in Table S1. Unvaccinated participants were on average younger, more likely to be non-White, less educated, and more likely to be female. Reported reasons for not being vaccinated are also summarized in Table S1, with the most common reason being concern over long-term health impacts from the vaccine.

### Vaccination side effects and vaccine type

When comparing dose 1 of the Moderna vaccine to dose 1 of the Pfizer-BioNTech, results showed participants receiving the Moderna vaccine had higher odds of having body ache, fatigue, fever/chills, nausea, pain at the injection site, redress at the injection site, and an overall larger number of reported side-effects (Figure 1a). When comparing dose 2 of the Moderna vaccine to dose 2 of the Pfizer-BioNTech vaccine, results showed participants receiving the Moderna vaccine had higher odds of having abdominal pain, body ache, fatigue, fever/chills, headache, nausea pain at the injection site, redness at the injection site, and an overall larger number of reported side-effects. When comparing the single-dose Johnson & Johnson vaccine to dose 2 of the Pfizer-BioNTech vaccine, results showed that those receiving the Johnson & Johnson vaccine had higher odds of body ache, fever/chills, headache, and overall total number of reported side effects. Johnson & Johnson subjects had lower odds of pain at the injection site and redness at the injection site compared to Pfizer-BioNtech (Figure 1b).

**Figure 1.**
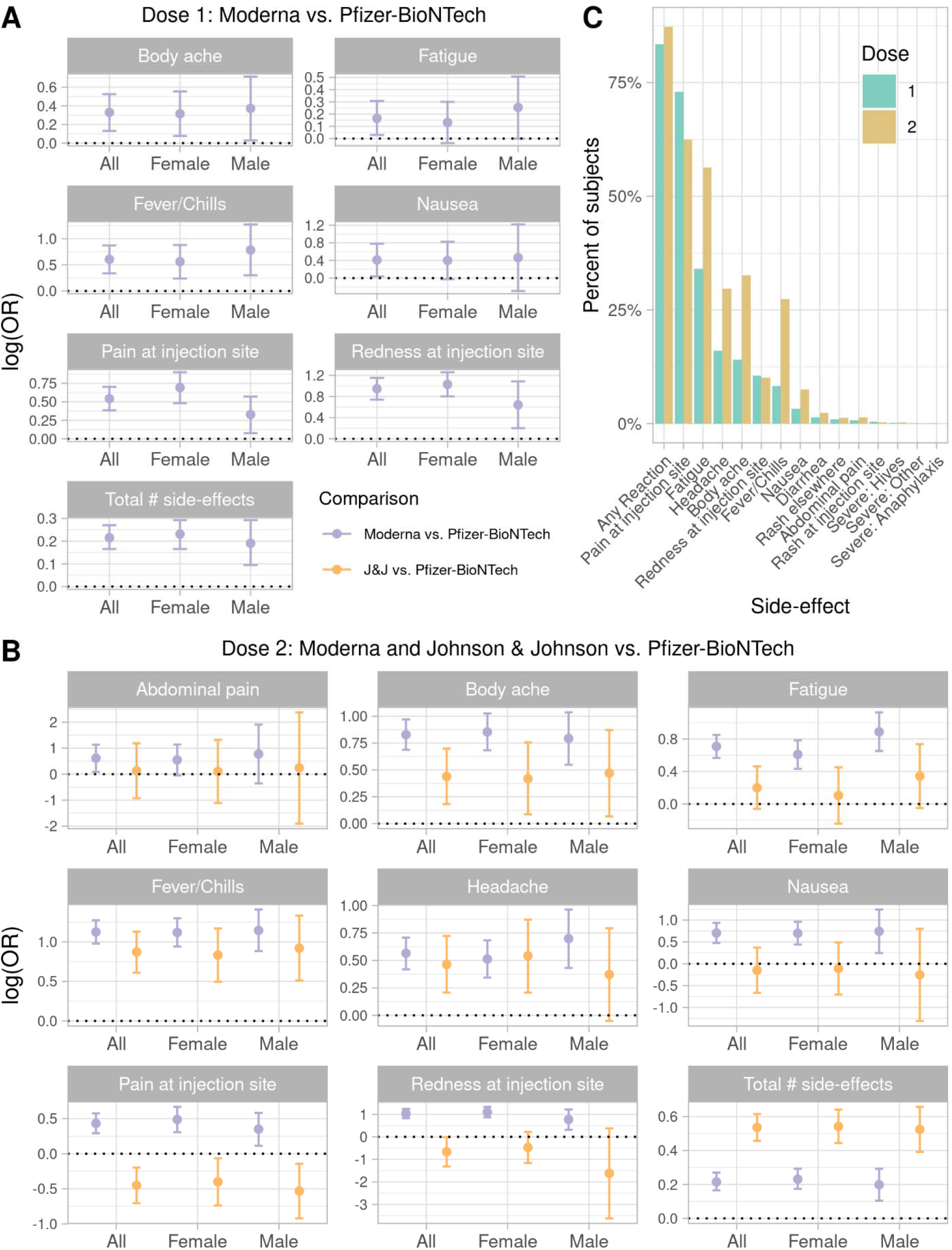
Vaccination side-effects. (A) Side-effect comparison of Moderna vs Pfizer-BioNTech Dose 1, (B) Side-effect comparison between Moderna and Johnson & Johnson vs Pfizer-BioNTech Dose 2, and (C) Percent of subjects reporting individual side-effects after Dose 1 and Dose 2.

### Vaccination antibody response and side effects

Frequencies of reported side-effects after dose 1 and dose 2 of vaccinations can be found in Figure 1c. Differences in anti-Spike antibody response (measured using S/C ratio) by presence or absence of reported side effects are summarized in Table 2. Side-effects after dose one of each vaccine were not significantly associated with antibody response. After dose 2, for each additional reported side-effect, subjects’ S/C ratio increased by 10.4%. Specifically, reporting fever/chills, headache, fatigue, or body ache after the second vaccination was associated with a 51.8%, 27.3%, 25.9%, or 33.4% increase in S/C ratio, respectively.

**Table 2.** Differences in antibody response (measured by S/C ratio) by presence or absence of reported side-effect.

|  | Difference in S/C % (95% CI) |  |
| --- | --- | --- |
|  | > 2 weeks after 1<sup>st</sup> dose | > 2 weeks after 2<sup>nd</sup> dose |
| **Demographics** |  |  |
| Age |  |  |
| Reference: <65 | 0 | 0 |
| ≥65 | -44.3 (-30.3, -55.6) | -24.1 (-0.2, -42.3) |
| Sex |  |  |
| Reference: Male | 0 | 0 |
| Female | +31.0 (5.7, 62.5) | +42.8 (7.3, 90.2) |
| Each additional day since vaccination | +6.9 (4.8, 9.0) | +0.4 (-1.5, 0.7) |
| **Side-effects** |  |  |
| Total No. side effects | +11.1 (-2.7, 13.3) | +10.4 (4.1, 17.1) |
| Fever/chills | +32.1 (-12.6, 99.5) | +51.8 (22.1, 88.6) |
| Headache | +5.7 (-23.0, 45.3) | +27.3 (2.1, 58.8) |
| Fatigue | +11.7 (-11.7, 41.2) | +25.9 (2.5, 54.7) |
| Body ache | +16.5 (-15.3, 60.3) | +33.4 (8.5, 64.1) |
| Nausea | +34.5 (-36.9, 186.6) | +13.8 (-21.2, 64.4) |
| Pain at injection site | +8.1 (-13.9, 35.6) | -0.8 (-19.0, 21.5) |
| Redness at injection site | +3.8 (-22.8, 39.5) | +10.4 (-17.5, 47.8) |
| **Vaccine type** |  |  |
| Reference- Moderna | 0 | 0 |
| Pfizer-BioNTech | -18.1(-34.5, +2.5) | -72.4 (-66.1, -77.5) |

### Vaccination antibody response and age and sex

Differences in antibody response (measured by S/C ratio) by age and sex are described in Table 2 and Figure 2a and 2b. For subjects who had only received the first dose of their vaccination and had more than two weeks between their blood sample and vaccination, there was a 44.3% decrease in S/C ratio for those age 65 and older. Females in this subgroup had a 31% greater S/C ratio compared to males. Additionally, for each additional day between vaccination and blood sample, S/C ratio increased by 6.9%. For subjects who were more than two weeks past their second dose of the vaccine, the only difference was a 42.8% larger S/C ratio in females compared to males.

**Figure 2.**
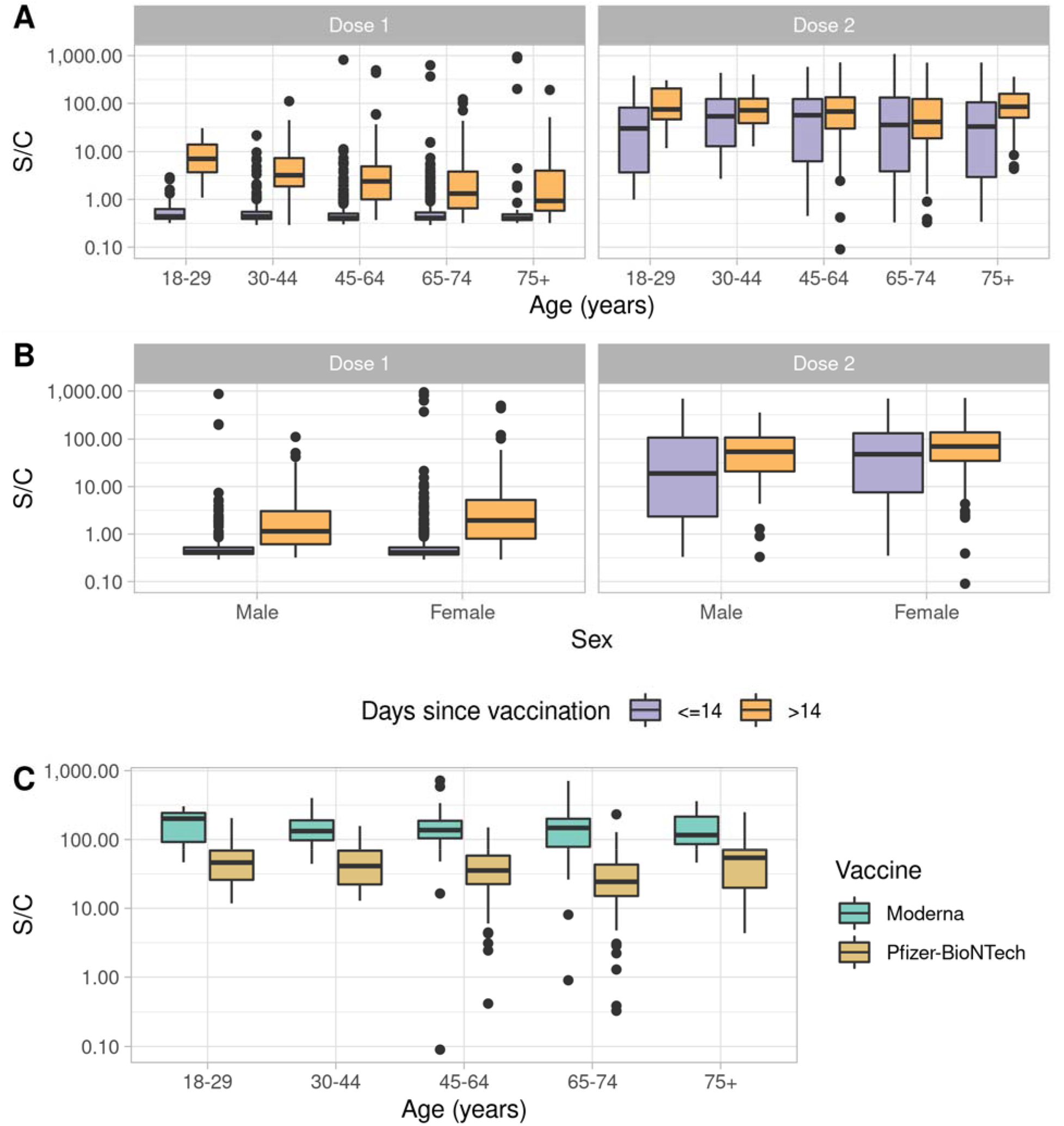
Vaccination antibody response. (A) Antibody response by age before and after 14 days post-vaccination. (B) Antibody response by sex before and after 14 days post-vaccination. (C) Difference in fully vaccinated antibody response by age between Moderna and Pfizer-BioNTech.

### Vaccination antibody response and vaccine type

Antibody response also differed between Moderna and Pfizer-BioNTech. At all vaccination timepoints (anyone with dose 1, more than two weeks after dose 1, anyone with dose 2, and more than two weeks after dose 2) Moderna recipients had a higher S/C ratio. Most importantly, those who were fully vaccinated with Pfizer-BioNTech had a 72.4% lower S/C ratio compared to those fully vaccinated with Moderna, adjusting for age, sex, and the number of days between vaccination and blood collection (Table 2, Figure 2c).

### Vaccination antibody response and other health factors

Additionally, antibody response differed between subjects reporting pre-existing health conditions compared to those that did not. Frequencies of these conditions can be found in Table 3. After adjusting for age, sex, vaccine type, vaccine dose, and number of days between vaccination and blood collection, subjects with asthma, diabetes, and cardiovascular disease all showed more than a 20% decrease in S/C ratio compared to those without these conditions (Table 3).

**Table 3.**
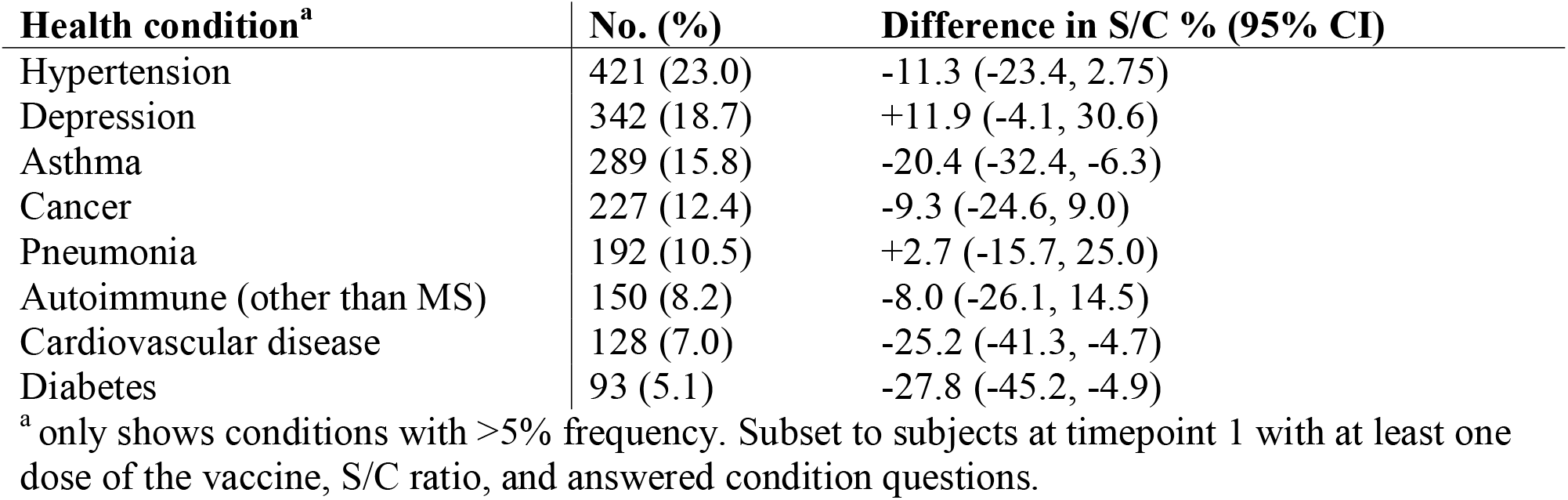
Prevalence and relationship between health conditions and serostatus in vaccinated participants at timepoint 1.

| **Health condition<sup>a</sup>** | **No. (%)** | **Difference in S/C % (95% CI)** |
| --- | --- | --- |
| Hypertension | 421 (23.0) | -11.3 (-23.4, 2.75) |
| Depression | 342 (18.7) | +11.9 (-4.1, 30.6) |
| Asthma | 289 (15.8) | -20.4 (-32.4, -6.3) |
| Cancer | 227 (12.4) | -9.3 (-24.6, 9.0) |
| Pneumonia | 192 (10.5) | +2.7 (-15.7, 25.0) |
| Autoimmune (other than MS) | 150 (8.2) | -8.0 (-26.1, 14.5) |
| Cardiovascular disease | 128 (7.0) | -25.2 (-41.3, -4.7) |
| Diabetes | 93 (5.1) | -27.8 (-45.2, -4.9) |

<sup>a</sup> only shows conditions with >5% frequency. Subset to subjects at timepoint 1 with at least one dose of the vaccine, S/C ratio, and answered condition questions.

## Discussion

As the COVID-19 pandemic and vaccination efforts continue, it is important to fully understand factors that influence vaccine response in representative cohorts. For participants in the East Bay COVID-19 Study, nearly all subjects were vaccinated by the second timepoint of questionnaire biospecimen collection in a context of very low natural SARS-CoV-2 infection (2.18%)^15, 16^. Subjects reported on both local and systemic vaccination side-effects, with ∼80% of subjects reporting at least one side-effect due to vaccination. The most common side-effects were pain at the injection site and fatigue. We found a higher antibody response to the Moderna vaccine in comparison to the Pfizer-BioNTech vaccine, which aligns with findings from previous studies^6-10^. Importantly, the majority of these previous studies either had a small sample size or were conducted in Europe where the AstraZeneca vaccine was widely administered. Our results show similar findings in a large and well-characterized cohort where the majority of participants received the two most widely administered mRNA vaccines, Moderna and Pfizer.

An important finding of this study was that increased side-effects were associated with increased antibody response two weeks after the second dose of the Moderna and Pfizer-BioNTech vaccine, providing evidence that a stronger reaction to the vaccine may confer greater protection against COVID-19.

Published studies have shown that immunocompromised patients have a weakened immune response^14^ and are more likely to have a breakthrough infection^17^. We observed that subjects with asthma, cardiovascular disease, and diabetes also demonstrated a weaker immune response (lower vaccine-induced antibody responses) compared to subjects without these pre-existing health conditions. Given that these conditions are also risk factors for more severe COVID-19 disease course, it is important to consider these conditions as we prioritize subjects for subsequent booster vaccination doses. These results persisted after adjustment for age, sex, and vaccine-related factors, including vaccine type, doses received, and lag between vaccination and biospecimen collection, increasing confidence that these results are not due to external factors. These conditions are known to impact the immune system and immunity. Studies have shown associations between primary immunodeficiencies and asthma and that people with asthma are at increased risk of respiratory infections and have a lower interferon response upon infection^18^. Other studies have found that the immune system plays a role in the onset of cardiovascular disease and that higher activity of the innate immune system and lower activity of the adaptive immune system may increase risk of cardiovascular disease^19^. Type 2 diabetes is characterized by chronic inflammation, and hyperglycemia can cause a dysfunctional immune response in patients that can lead to increased susceptibility to infection^20^.

A major strength of our study is the design, which includes a large and well-characterized cohort of ∼5,000 individuals with high vaccination rates and biospecimen sample collection from subjects at all stages of vaccination (<2 weeks, 1 dose; >2 weeks, 1 dose; <2 weeks, 2 dose; fully vaccinated). Side-effects were assessed after each vaccination dose for all participants. Due to the end date and short-term nature of this study, we were unable to assess long-term antibody measurements or booster shot responses. We also have no reports on break-through infections after vaccination, although it is important to note that infection rates were extremely low in this cohort prior to vaccination. Because of the low infection rates, we also do not have a large enough sample size of previously infected individuals to look at differences in vaccination response between previously infected and uninfected subjects.

## Conclusion

The findings of this study of a well-characterized population-based cohort in the East Bay of Northern California show lower antibody responses for those receiving the Pfizer-BioNTech vaccine compared to the Moderna vaccine. Additionally, we report lower antibody responses for subjects with pre-existing conditions that are risk factors for COVID-19 severity, including asthma, diabetes, and cardiovascular disease. These findings are important to consider as people with these pre-existing conditions are considered along with the overall public for future vaccination booster doses.

## Data Availability

De-identified individual participant data that underlie the results reported in this article (text, tables, figures may be shared for up to 36 months following publication after investigators whose proposed use of the data has been approved by an independent review committee. For individual participant data analysis or meta-analysis, proposals should be directed to Lisa Barcellos and Lynn Hollyer. Requests will be reviewed by an independent review committee and the UC Berkeley Institutional Review Board. Data may be shared upon approval. Data requestors will need to sign a data access agreement.

## Acknowledgements

We want to acknowledge the contributions of the following individuals to this study: Ella Parsons, Jordan Keen, Janine Solomon, Jose Salinas, Kevin Duong, Joseph Egbunikeokye, Maya Talavera, Riya Shrestha, Colin Warnes, Jose’ Victor Zambrana, Magelda Montoya, Nicholas Lo, Parnal Narvekar, Fausto Bustos, Gregorio Dias, Reinaldo Mercado-Hernandez, Julia Huffaker, Raymond Montes, Alexandra Zermeno, Alejandra Zeiger, William Dow, Michael Lu, Lila Krop, Kelly Lam, Yan Zhang, Sarah Folkmanis, Sophie Zhai, Dingjun Chen, Ruben Vargas Ethan Garcia, Oliver Li, Manisha Sahoo, Raina Walencewicz, Sophia Wang, Antonia Gibbs, Amrita Ramanathan, Catherine Livelo, Taylor Worley, Amanda Tanaka, Savinnie Ho, Jane Liu Ryan Allen, Sofia Soltero, Victoria Van Metter, Madeleine Fraix, Allie Coyne, Subeksha Sharma Lydia Yu, Shreeya Garg, Sanjeet Paluru, Malika Saxena, Talia Panadero, Ayra Rahman, Joshua Calangian, Dharaa Upadhyaya, Sophia Kemp, Ruhi Parikh, Amy Rich, Sophie Manoukian, Nola Vu, Crystal Nguyen, Jordyn Pinochi, Alma Kuc, Siri Ylenduri, Manvir Kaur, Angikaar Chana, and Sannidhi Sarvadhavabhatla, Benjamin T. Auch, Dinesha Walek, Evan Forsberg, Jerry Daniel, Veronica Tonnell, Ji Hyun (Jay) Kim, Mary Nieuwenhuis. Creative Testing Solutions: Valerie Green, Sherri Cyrus, Phillip Willamson, Brett Hirsch, Paul Contestable, Mars Stone, Joe Derisi, Emily Crawford, Emily Ahlvin, Armando Diaz, and Favianna Rodriquez.

---

**Table S1.**
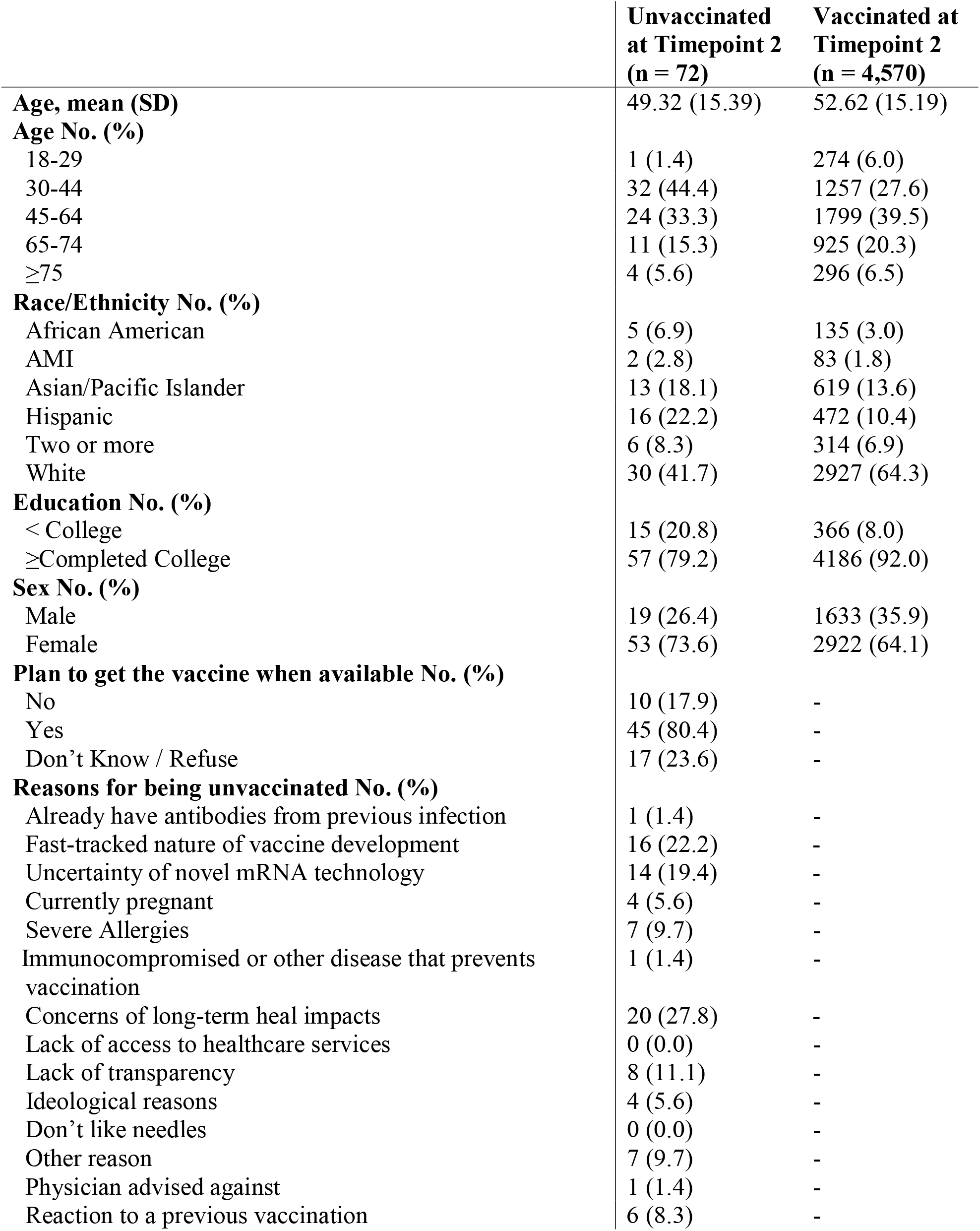
Characteristics of unvaccinated participants.

