## Supplemental Table 1 for "COVID-19 vaccine antibody response is associated with side-effects, chronic health conditions, and vaccine type in a large Northern California cohort"

|  | **Unvaccinated at Timepoint 2 (n = 72)** | **Vaccinated at Timepoint 2 (n = 4,570)** |
| --- | --- | --- |
| **Age, mean (SD)** | 49.32 (15.39) | 52.62 (15.19) |
| **Age No. (%)** |  |  |
| 18-29 | 1 (1.4) | 274 (6.0) |
| 30-44 | 32 (44.4) | 1257 (27.6) |
| 45-64 | 24 (33.3) | 1799 (39.5) |
| 65-74 | 11 (15.3) | 925 (20.3) |
| ≥75 | 4 (5.6) | 296 (6.5) |
| **Race/Ethnicity No. (%)** |  |  |
| African American | 5 (6.9) | 135 (3.0) |
| AMI | 2 (2.8) | 83 (1.8) |
| Asian/Pacific Islander | 13 (18.1) | 619 (13.6) |
| Hispanic | 16 (22.2) | 472 (10.4) |
| Two or more | 6 (8.3) | 314 (6.9) |
| White | 30 (41.7) | 2927 (64.3) |
| **Education No. (%)** |  |  |
| < College | 15 (20.8) | 366 (8.0) |
| ≥Completed College | 57 (79.2) | 4186 (92.0) |
| **Sex No. (%)** |  |  |
| Male | 19 (26.4) | 1633 (35.9) |
| Female | 53 (73.6) | 2922 (64.1) |
| **Plan to get the vaccine when available No. (%)** |  |  |
| No | 10 (17.9) | - |
| Yes | 45 (80.4) | - |
| Don’t Know / Refuse | 17 (23.6) | - |
| **Reasons for being unvaccinated No. (%)** |  |  |
| Already have antibodies from previous infection | 1 (1.4) | - |
| Fast-tracked nature of vaccine development | 16 (22.2) | - |
| Uncertainty of novel mRNA technology | 14 (19.4) | - |
| Currently pregnant | 4 (5.6) | - |
| Severe Allergies | 7 (9.7) | - |
| Immunocompromised or other disease that prevents  vaccination | 1 (1.4) | - |
| Concerns of long-term heal impacts | 20 (27.8) | - |
| Lack of access to healthcare services | 0 (0.0) | - |
| Lack of transparency | 8 (11.1) | - |
| Ideological reasons | 4 (5.6) | - |
| Don’t like needles | 0 (0.0) | - |
| Other reason | 7 (9.7) | - |
| Physician advised against | 1 (1.4) | - |
| Reaction to a previous vaccination | 6 (8.3) | - |

**Table S1. Characteristics of unvaccinated participants.**
